# Point-of-care serodiagnostic test for early-stage Lyme disease using a multiplexed paper-based immunoassay and machine learning

**DOI:** 10.1101/19009423

**Authors:** Hyou-Arm Joung, Zachary S. Ballard, Jing Wu, Derek K. Tseng, Hailemariam Teshome, Linghao Zhang, Elizabeth J Horn, Paul M. Arnaboldi, Raymond J. Dattwyler, Omai B. Garner, Dino Di Carlo, Aydogan Ozcan

## Abstract

Caused by the tick-borne spirochete, *Borrelia burgdorferi*, Lyme disease (LD) is the most common vector-borne infectious disease in North America and Europe. Though timely diagnosis and treatment are effective in preventing disease progression, current tests are insensitive in early-stage LD, with a sensitivity <50%. Additionally, the serological testing currently recommended by the US Center for Disease Control has high costs (>$400/test) and extended sample-to-answer timelines (>24 hours). To address these challenges, we created a cost-effective and rapid point-of-care (POC) test for early-stage LD that assays for antibodies specific to seven *Borrelia* antigens and a synthetic peptide in a paper-based multiplexed vertical flow assay (xVFA). We trained a deep learning-based diagnostic algorithm to select an optimal subset of antigen/peptide targets, and then blindly-tested our xVFA using human samples (*N*_(+)_ = 42, *N*_(−)_= 54), achieving an area-under-the-curve (AUC), sensitivity, and specificity of 0.950, 90.5%, and 87.0% respectively, outperforming previous LD POC tests. With batch-specific standardization and threshold tuning, the specificity of our blind-testing performance improved to 96.3%, with an AUC and sensitivity of 0.963 and 85.7%, respectively.

## INTRODUCTION

Lyme disease (LD) is the most common vector-borne infectious disease in both North America and Europe, causing ∼300,000 infections annually in the United States^1,2^. It is caused by infection with the spirochete *Borrelia burgdorferi* (Bb) transmitted by black-legged ticks (*Ixodes genus)*. Early disease is associated with a characteristic skin lesion, erythema migrans (EM) along with other symptoms^3–5^. If not diagnosed and treated with appropriate antibiotics, the infection can disseminate to distal sites including the nervous system, heart, and joints causing an array of symptoms, including e.g., lymphocytic meningitis, cranial neuropathy, facial nerve palsy, radiculopathy, A/V node heart block ^3,6,7^, and arthritis^4,8^.

Although a presenting EM is diagnostic, the characteristic lesion is absent in 10-20% of infected persons and is frequently atypical thus escaping recognition. This makes laboratory testing critical to confirm the diagnosis and guide treatment^3,5,6,9^ Despite recent advances in direct detection of Bb through e.g. Nucleic Acid Amplification Testing (NAAT), these methods remain inadequate due to the low concentration and transient presence of Bb in the blood. Culturing Bb is also not practical due the slow growth of the bacteria, as well as the need for specialized growth media ^9–11^. Therefore, current testing methods work indirectly by detecting specific antibodies produced by the body’s immune response to the infection.

The United States Center for Disease Control and Prevention (CDC) recommends a ‘two-tier’ testing method, where the first-tier consists of a sensitive Enzyme Immunoassay (EIA) or immunofluorescence assay (IFA). If the first-tier is positive or equivocal, a Western Blot (WB) is then recommended for confirming the presence of 2 of 3 Immunoglobulin M (IgM) antibodies and/or 5 of 10 Immunoglobulin G (IgG) antibodies targeting Bb associated antigens^12^. A number of reports have also showed the efficacy of a modified two-tier test (MTTT) format, where the WB is replaced by a second complimentary EIA, and as a result, the FDA has recently approved the use of some EIAs as viable tests for the second tier^13–17^.

Despite being the standard for the laboratory diagnosis of LD, the two-tier serological testing method has multiple drawbacks. Although there is a high specificity (>98%) and sensitivity (70-100%) in *late* LD, the two-tier test has poor sensitivity in *early-stage* LD, seldom exceeding 50% at the time when most patients seek medical care^6,9,14,18,19^. This is also the time when treatment is the least costly and most effective at preventing disease sequela. The poor sensitivity can be attributed to the underdeveloped immune response within the first weeks of infection in which a limited IgM antibody response is followed by an IgG antibody response. However, it is also exacerbated by the limited number of antigen-targets in the first tier test that may miss detection of antibodies produced during the earliest stage of infection.^20^ Specifically, the earliest responses are to Flagellin B (FlaB) and p66 with responses to a number of additional antigens such as OspC (25kd), VlsE, BBK32, FlaA (37kd), BmpA (39kd) and DbpA proteins developing as *B. burgdorferi* disseminates^20–23^.

The two-tier testing method also suffers from slow turn-around time (>24 hours) and high costs (>$400/test), with estimated expenses exceeding $492 million *annually* just in the United States^24^. Additionally, the standard testing used in the two-tier format must be performed in a centralized testing facility by trained technicians, requiring bulky and expensive clinical analyzers. These drawbacks therefore limit accessibility to accurate LD testing, especially impacting populations far from clinical laboratories as well as populations in rural and forested areas where tick bites are common. Therefore, accurate and affordable LD testing methods for use at the point-of-care (POC) are in high demand^13,18^.

Paper-based lateral flow assays (LFAs), also known as Rapid Diagnostic Tests (RDTs), are appealing for POC serological analysis due to their low-cost, ease-of-use, and rapid nature^25–27^. These tests use e.g., colorimetric or fluorescent conjugates embedded in one-time-use cassettes to rapidly and cost-effectively detect the presence of antibodies specific to disease. LFAs however, are not conducive to the detection of multiple analytes due to their in-line geometry, measuring only one or two antibodies in a single test^28^.

This restrictive design inherently limits the potential sensitivity and specificity of traditional RDTs for LD. For example, tests that rely on single antibody measurements (like many EIAs in the first tier) can be less robust to false positive results due to antigens (p66 and FlaB in particular) which contain epitopes that are highly cross-reactive with epitopes found in multiple other bacteria^29,30^. They can also have low sensitivity if the target antigen is mismatched with the underlying immunodominance. This paradigm is the reason that the performance of the two-tier algorithm can depend on which EIA is used in the first tier, as well as what strain of Bb the EIA test was designed against, B31 being the most common^31^.

Therefore, to overcome this limitation, large-scale screening efforts alongside new epitope mapping and peptide synthesis are focused on developing a universal multi-antigen detection panel, with e.g. 5 to 10 LD-specific antigen targets being suggested for improving diagnostic performance for early LD^17,32,33^. Consequently, we have a unique opportunity to leverage advances in computation and machine learning, to train novel serodiagnostic algorithms with these rich, multi-antigen measurements derived from well-characterized clinical samples, to ultimately create new decision algorithms that outperform the traditional two-tier test^32–34^. Deep learning, which refers to the use of artificial neural networks with multiple hidden-layers, can be especially effective in developing nonlinear yet robust inference models from noisy data-sets with complex and confounding variables^35–37^.

Here, we introduce a paper-based multi-antigen POC test powered by deep learning for serological diagnosis of early-stage LD. Our test is a multiplexed vertical flow assay (xVFA), composed of a stack of functional paper layers, which in contrast to the more common LFA, allows for a multi-antigen detection panel for measuring an array of LD-specific antibodies on a single sensing membrane. Containing 13 spatially-separated immunoreaction spots, the sensing membrane is functionalized with Bb-specific antigens (OspC, BmpA, P41, DbpB, Crasp1, P35, Erpd/Arp37) as well as a peptide (Mod-C6) composed of a C6-like epitope linked to a specific p41 epitope. The xVFA can be operated in 15 min, after which the assay cassette is opened and the sensing membrane is imaged by a custom-designed mobile-phone based reader. Computational analysis then quantifies the colorimetric signals on the sensing membrane through automated image processing, and a neural network is used to automatically infer a diagnosis from the multiplexed immunoreactions. The diagnostic algorithm in this work was trained with 50 human serum samples (25 early-stage LD and 25 endemic controls), obtained from the Lyme Disease Biobank (LDB), run in duplicate for both IgM and IgG antibodies, resulting in 200 individually-activated xVFAs composing the training data-set. This training data-set was also used to computationally select a subset of detection antigens from the larger panel using a feature selection technique, improving the diagnostic performance and reducing the per-test cost. The computational xVFA was evaluated through blind testing of an additional 50 human serum samples (25 early stage LD and 25 endemic controls), obtained fully-blinded by the LDB. Testing entirely early-stage LD samples, we achieved an AUC of 0.95, and by equally weighing the false positive and false negative results, we obtained a sensitivity and specificity of 90.5% and 87.0% with respect to the gold-standard two-tier serological testing. By adjusting the diagnostic cut-off value to favor high-specificity during the training phase and incorporating batch-specific standardization for the network inputs, our blind testing specificity improved to 96.3%, with a small drop in our sensitivity (85.7%) in relation to the gold-standard two-tier serological testing.

There is currently a *first tier* POC LD test, but *no* FDA-approved POC test for LD that can serve as a replacement to the two-tier method^28,38^. However, development of a POC two-tier replacement test will allow for more rapid diagnosis and better treatment outcomes. This is especially important as LD is projected to increase over the next decades as the geographic areas of tick-populations continue to expand ^39–41^. Although multi-target POC sensing approaches for LD have been explored in the literature, the methods proposed either exhibited poor performance or have not undergone validation with a clinical study^28,42^. We believe our platform demonstrates a leapfrog improvement over existing POC LD testing approaches, reporting a cost-effective and rapid (15 min) paper-based multiplexed assay powered by deep learning for serological diagnosis of early-stage LD at the POC.

## MATERIALS AND METHODS

### The multiplexed vertical flow assay (xVFA)

#### Overview

Our multi-target vertical flow assay (xVFA) is composed of a stack of functional paper layers and a sensing membrane contained within a 3-D printed plastic cassette. The cassette is divided into a top and bottom-case which can be separated through a twisting mechanism, revealing the multiplexed sensing membrane on the top layer of the bottom-case. The sensing membrane contains 13 immunoreaction spots defined by a black wax-printed barrier, where each spot is pre-loaded with a different capture-antigen or antigen epitope-containing peptide as well as proteins serving as positive- and negative-controls to enable multiplexed sensing information within a single test (Fig. 1B). For each xVFA, there are two top-cases used during the operation (Fig. 1C). The first top-case facilitates the uniform flow of a serum sample from the loading inlet to the sensing membrane, where LD-specific antibodies are bound to the detection antigens immobilized on the nitrocellulose surface. The second top-case is then used for color signal generation, where a conjugate pad, upon wetting, releases embedded gold-nanoparticles (AuNPs) conjugated to anti-human IgM or IgG antibodies. The AuNPs then bind to the LD-specific IgM or IgG antibodies previously captured on the sensing membrane, resulting in a color signal in response to the captured amount. After completion of these sandwich immunoreactions, both IgM and IgG running in parallel, the sensing membrane is immediately imaged by a custom-designed mobile-phone reader (Fig. 1D-E), which captures the background image (taken before the assay operation) and the signal image (taken after the assay operation) of the sensing membrane for subsequent analysis in a computer, where a neural network is used to ultimately determine the final result (seropositive or seronegative). The general concept of paper-based vertical flow design was reported in our previous work^43^; a detailed breakdown of the functional paper layers and assay reagents can be found in the Supplementary Information (Fig. S1, Table S1, S2) along with further discussion on the wax-printed sensing membrane design and optimized operational protocol (Fig. S2).

**Figure 1.**
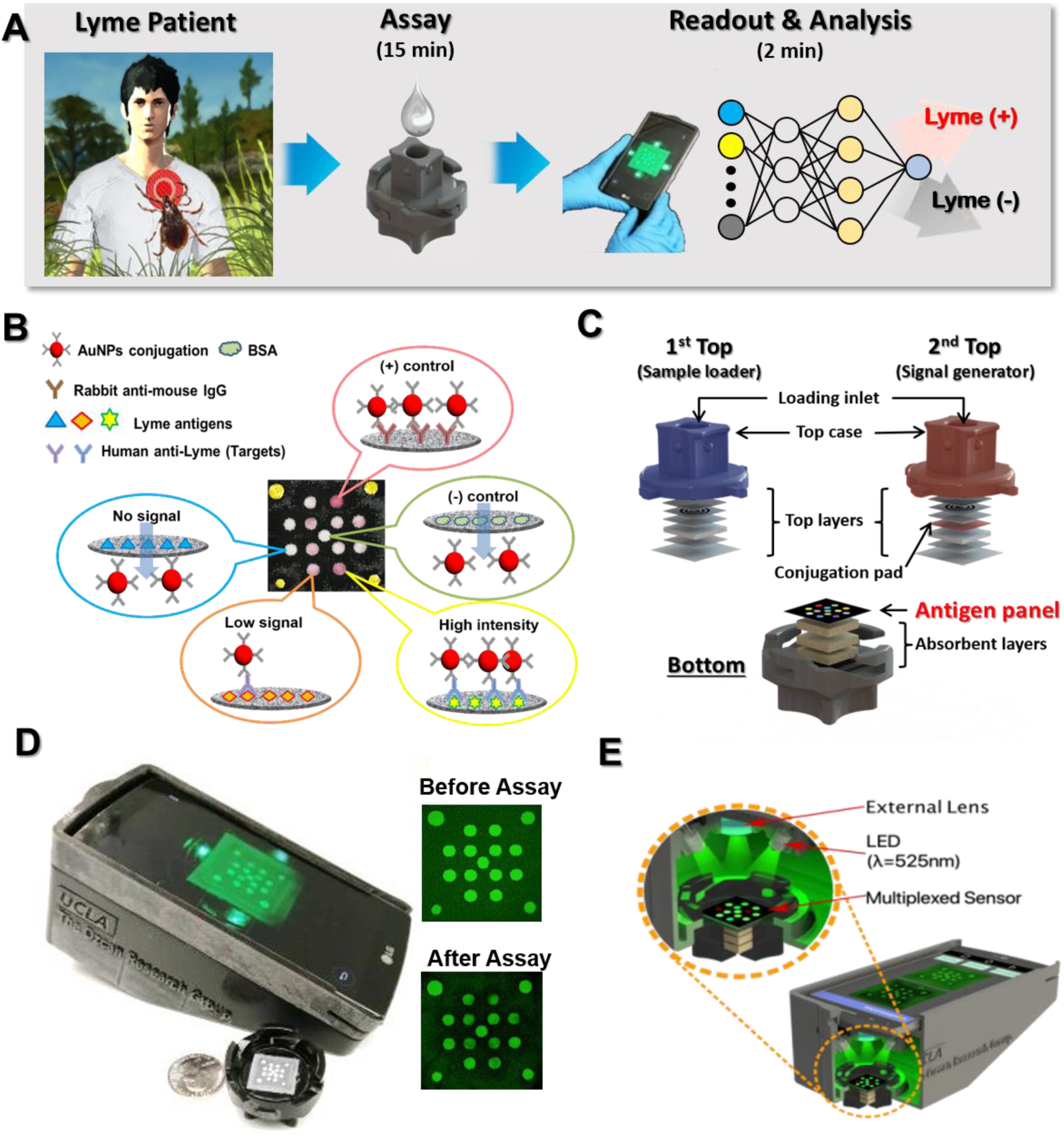
**(A)** Overview of POC Lyme disease diagnostic testing using xVFA and machine learning. **(B)** Illustration of the multiplexed immunoreactions which occur on the sensing membrane during the xVFA operation. **(C)** Expanded diagram of the paper materials within the xVFA showing the sample loading top-case (in blue) and the signal generating top-case (in red). The bottom-case is shown below with the sensing membrane containing the multi-antigen panel. **(D)** Photograph of the mobile-phone reader with an opened xVFA cassette and example images of the sensing membrane (shown to the right). **(E)** Cross-section of the xVFA mobile-phone reader with an inset showing the illumination of the sensing membrane using an inexpensive opto-mechanical attachment to the smartphone.

The mobile phone-based assay reader (Fig. 1D-E) is constructed from a smartphone (LG G4H810) and 3-D printed (Dimension Elite, Stratasys) attachment containing four 525 nm wavelength light emitting diodes (LEDs) for even illumination of the sensing membrane. An external lens was also mounted in the 3-D printed attachment below the built-in phone camera lens system for enabling an in-focus field of view. All the images were obtained in raw dng format using the standard Android camera app of the smartphone.

#### Assay operation

First, a background image of the blank sensing membrane is taken with our mobile-phone reader. Then the first top-case is mated with the bottom-case, and 200 µL of running buffer is introduced to fully wet the paper layers in the xVFA. After the buffer is absorbed fully into the xVFA cassette (∼20 seconds), 20 µL of serum sample is pipetted into the loading inlet and allowed to absorb. Then, a second addition of running buffer is introduced to the loading inlet, followed by a 6-minute wait period, during which the serum sample reacts with the sensing membrane and the unreacted sample is washed away to the lower absorbent pads. The first top-case is then exchanged with the second top-case, and 450 µL of running buffer is added to release the AuNP conjugates responsible for color signal generation. After an 8-minute wait period, the xVFA cassette is opened and imaged by the mobile-phone reader to get the multiplexed signal.

Separate xVFAs are run in parallel for IgM and IgG antibody detection, where the only difference between the two xVFAs is in the conjugate pad, i.e. one conjugate pad contains AuNPs conjugated to anti-IgM antibodies and the other contains AuNPs conjugated to anti-IgG antibodies (Fig. 2A).

**Figure 2.**
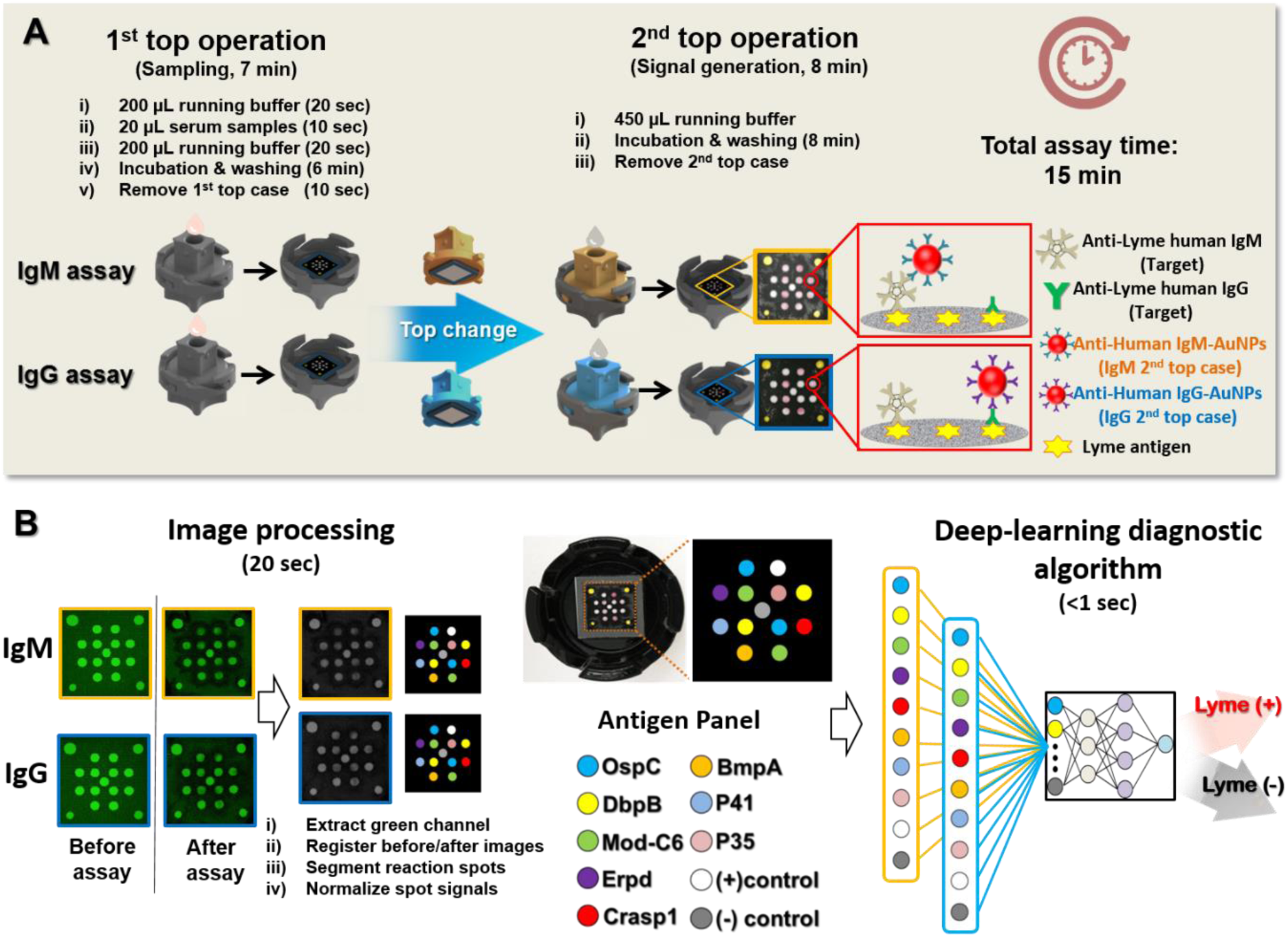
**(A)** Detailed xVFA operation for the IgM and IgG assays, which are performed in parallel (total assay time: 15 min). A depiction of the IgM and IgG immunoassays is shown to the right. **(B)** Image processing (left), capture antigen panel (middle), and deep learning-based analysis (right) of the multiplexed sensing membrane.

#### Image processing and deep learning-based analysis

Raw dng images, captured by our mobile reader, of the sensing membrane taken before (background image) and after (signal image) the assay are converted to tiff format. The green pixels are then extracted, and the background and signal images are registered to each other via a rigid-transformation. The immunoreaction spots are then identified in the background image and a fixed-radius mask is defined per-spot at approximately 80% of the immunoreaction spot area. The pixel intensity within this immunoreaction spot mask is then calculated for the registered signal image and normalized by the pixel average of the corresponding immunoreaction spot in the registered background image,

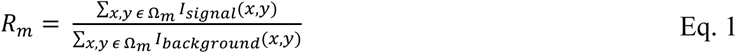

where *R*_*m*_ defines the normalized signal per immunoreaction spot (*m*) and Ω_*m*_ defines the x-y bounds of the fixed-radius mask per immunoreaction spot. This background normalization procedure helps account for non-uniformities in the illumination as well as local defects that might exist within the immunoreaction spots on each xVFA sensing membrane. Immunoreaction spots functionalized by the same capture antigen are averaged together, and each of the unique *R*_*m*_ signals, derived from both the IgM and IgG xVFAs, are then used for deep learning analysis (Fig. 2B).

Lastly, before being input into the diagnostic decision neural network, the *R*_*m*_ signals from both the IgM and IgG xVFAs are standardized to the mean, 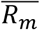, and standard deviation, *σ*_*m*_ taken over the *training* set,

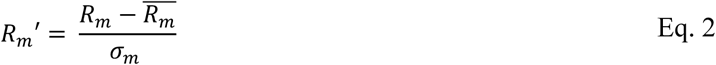

The decision neural network contains an input layer with M nodes (e.g., M = 20 with the full IgM/IgG antigen panel), three fully connected hidden layers with 128, 64, and 32 nodes, in the first, second, and third layers, respectively. Each layer contains batch normalization, a 50% dropout, and a Rectified Linear Unit (ReLU) activation function, defined by *f*(*x*) = max(0, *x*), with the exception of the final output layer, which uses a sigmoid activation function, yielding a network output as a numerical value between 0 and 1. A final binary diagnosis is then made by evaluating this numerical output with a blind cutoff value of 0.5. A binary cross-entropy cost function, defined as:

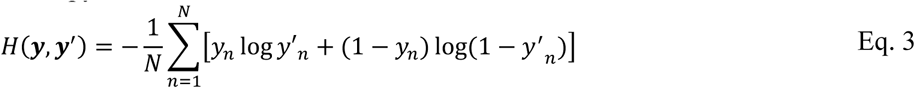

was used during the training phase (learning rate = 0.001, batch-size = 32) with the two-tier based seropositive and seronegative diagnosis used as the gold-standard label, *y*_*n*_ *ϵ* {0,1}. Here, *N* represents the number of training samples, and *y*′_*n*_ *ϵ* (0,1) represents the neural network prediction. The hyper-parameters mentioned above (except the diagnostic threshold) were determined by a random parameter optimization in which 2 and 3 hidden layers with varying number of nodes were tested via k-fold cross-validation (k =5) along with different dropout, learning rate, and batch size parameters randomly selected from a predefined list. All networks were trained for up to 1000 epochs with early stopping criterion defined by the stagnation of training accuracy not exceeding a change of 0.1% over 100 training epochs. The network weights at the last epoch (defined by the early stopping criterion) were then stored in the final model.

### Clinical study

In total, 106 unique human serum samples were obtained from the LDB (collected under Advarra IRB Pro00012408). Out of these sample, 50 were used for training, 50 for blind testing, with the additional 6 used for precision evaluation. All the samples used were *early-stage* LD, having been obtained < 30 days since symptoms or the initial tick bite. All the cases and endemic controls were confirmed to be Lyme-positive or negative through standard two-tier testing methods, or in some cases quantitative Polymerase Chain Reaction (qPCR) or convalescent draws (seroconversion). For the first tier, a combination of Whole Cell Lysate Enzyme-linked Immunosorbent Assay (ELISA), C6 Peptide EIA, or VlsE/PepC10 ELISA testing was used. The second tier, performed regardless of the first-tier results, was comprised of the standard IgM and IgG WB. For this work, samples were considered seropositive if any of the three EIA tests in the first tier had a positive or equivocal (borderline) result and the second tier had a positive result for either the IgM or IgG WB as defined by the CDC recommendation (≥ 2 of 3 bands for IgM WB, and ≥ 5 of 10 bands for IgG)^9^. Samples were also considered seropositive by MTTT guidelines, where a seropositive diagnosis was called from two positive or equivocal EIA tests along with the presence of EM, without the need for a positive WB result^16,31^. Additionally, all samples were confirmed negative for coinfections of Anaplasmosis and Babesia, both of which are infections also transmitted by the *Ixodes* tick and can produce similar constitutional symptoms to LD.

The clinical samples used for training and testing were obtained and tested in two separate sample-pulls. The first sample-pull contained 25 LD cases and 25 endemic controls from a collection site in East Hampton, New York between 2014 and 2016 (see Table S3). 24 of the 25 LD cases were seropositive, with the one exception confirmed Lyme-positive through *B. burgdorferi* qPCR. All seropositive samples were early-stage LD, with 3 out of 24 being early disseminated, defined by the presence of multiple EMs. All samples in the first sample-pull were tested with the IgM and IgG xVFAs in duplicate and used as the *training* data-set (*N*_(+)_ = 48, *N*_(−)_= 52). We should emphasize that since our *training ground truth* for this work is the two-tier test (which we aim to replace using our paper-based POC xVFA), one of the Lyme-positive samples here (tested in duplicate) has been added to the negative set (*N*_(−)_= 52) since it was seronegative (although being qPCR positive).

The second sample-pull, used for blind testing of our platform, also contained 25 LD cases and 25 endemic controls, but was obtained fully blinded (i.e. without case and control labels), and tested with our xVFAs and the associated serodiagnostic algorithm three months after the first sample-pull (see Table S4). The second sample-pull also contained cases and endemic controls from collection sites in East Hampton (USA), New York (USA), but also included samples from several sites in Wisconsin (USA), which were *not* included in our training phase. In this blind testing set, 23 of the 25 LD cases were seropositive, with the two exceptions confirmed Lyme-positive through a convalescent blood-draw taken approximately 3 months after the first draw that revealed seroconversion via two-tier testing methods. All seropositive samples were early-stage LD, with 9 out of 23 being early disseminated. All the samples in the second sample-pull were tested with the IgM and IgG xVFAs in duplicate and used as a blinded test set (*N*_(+)_ = 42, *N*_(−)_= 54), with four IgM tests removed due to fabrication error (see Supplementary Information, Fig. S3). The seropositive and negative diagnostic predictions resulting from our xVFA testing and machine learning algorithm were blindly sent to the LDB prior to receiving the gold-standard testing labels (to determine our blind testing performance). Six sensing membranes were additionally processed (compared to other activated xVFAs) using an affine image transformation to correct misalignments resulting from unforeseen expansion and/or contraction of the nitrocellulose pad, during the automated image registration step (see Supplementary Information, Fig. S4).

Lastly six separate serum samples, obtained at the same time as the second sample pull were used for precision evaluation performed by a single assay operator as well as multiple, newly trained personnel. In summary, 106 unique human serum samples obtained from the LDB have been used for training, testing and further inspection of our xVFA LD detection platform.

## RESULTS

### Training and cross-validation

The first sample-pull (*N*_(+)_ = 48, *N*_(−)_= 52) was used entirely for cross-validation and training of our xVFA platform. Figure 3 summarizes the signals from the multi-antigen detection panel across all the seropositive samples in the training set, with the raw (1 − *R*) signals of all the tests shown in Figure S5. The statistical performance of each antigen in terms of its t-score (Eq. S1) is also shown, with the Mod-C6 peptide and OspC antigen being the top-ranked immunoreaction spots for the IgG and IgM sensing membranes respectively. As will be detailed and discussed later on, the individual performance and therefore the value of different antigen-targets as a single disease discriminator is highly-limited in the context of LD diagnosis, and our multiplexed xVFA platform computationally selects a complementary set of antigen-targets that collectively diagnose early-stage LD, providing a cost-effective and rapid POC replacement for the two-tier test.

**Figure 3.**
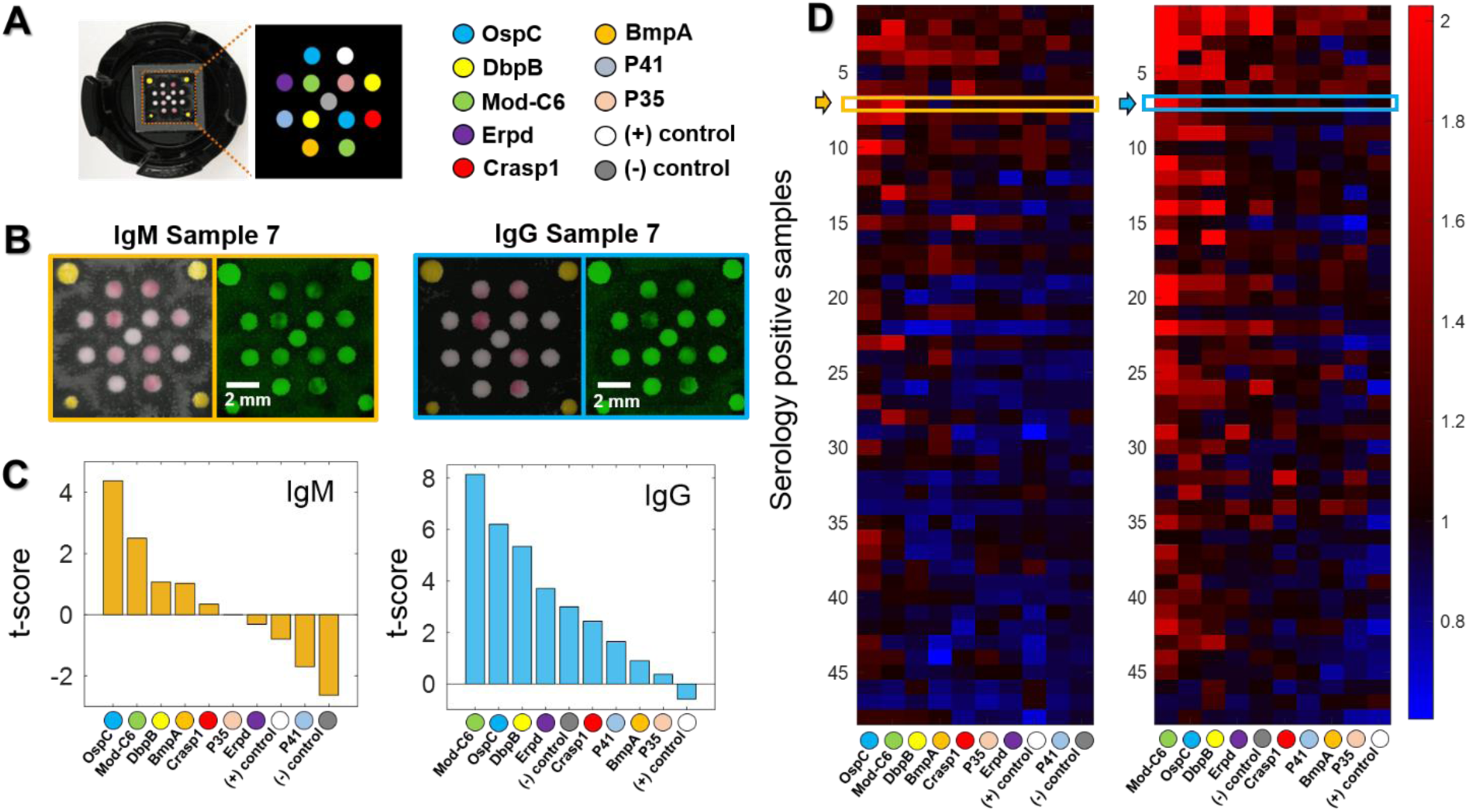
**(A)** The sensing membrane of our xVFA and the map of the multi-antigen panel. **(B)** Example images of IgM (left) and IgG (right) sensing membranes after activated by a human serum sample (sample #7) under ambient lighting conditions and using the mobile-phone reader (under green LED illumination). **(C)** The t-score (Eq. S1) of the multi-antigen channel calculated over the training data-set for the IgM (left) and IgG (right) sensing membranes ranked in descending order. **(D)** A heat map showing the signals from the multi-antigen panel of the IgM (left) and IgG (right) sensing membranes activated by the seropositive samples in the *training* set. The color bar represents the (1-R) signal of each sensing channel normalized to the mean signal of the same sensing channel across all the seronegative samples in the training data-set. The normalized IgM and IgG signals for sample #7 are outlined by the orange and blue boxes respectively.

Before training the final serodiagnostic algorithm to be used for the detection of early-stage LD, the training set was used for selecting an optimal subset of antigens. We implemented a sequential forward feature selection (SFFS) method where the signals from each sensing channel were added one at a time, into the input layer of the neural network and then trained via k-fold (k=5) cross-validation. After the addition of each input feature, the performance of the network was evaluated based off a mean square error cost function, i.e.,

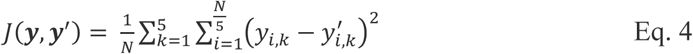

where *y*_*i,k*_ *ϵ* {0,1} is the ground truth binary result (*i*.*e*. seropositive or negative) and 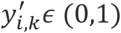 is the numerical output of the network for the i^th^ test in the k^th^ fold of the k-fold cross validation (k=5). *N* is the number of tests in the training data-set. The input feature which yielded the best network performance for that iteration was then kept as an input feature until all the 20 sensing channels were included as inputs (for ranking of their collective value for LD diagnosis); see e.g., Fig. 4C.

**Figure 4.**
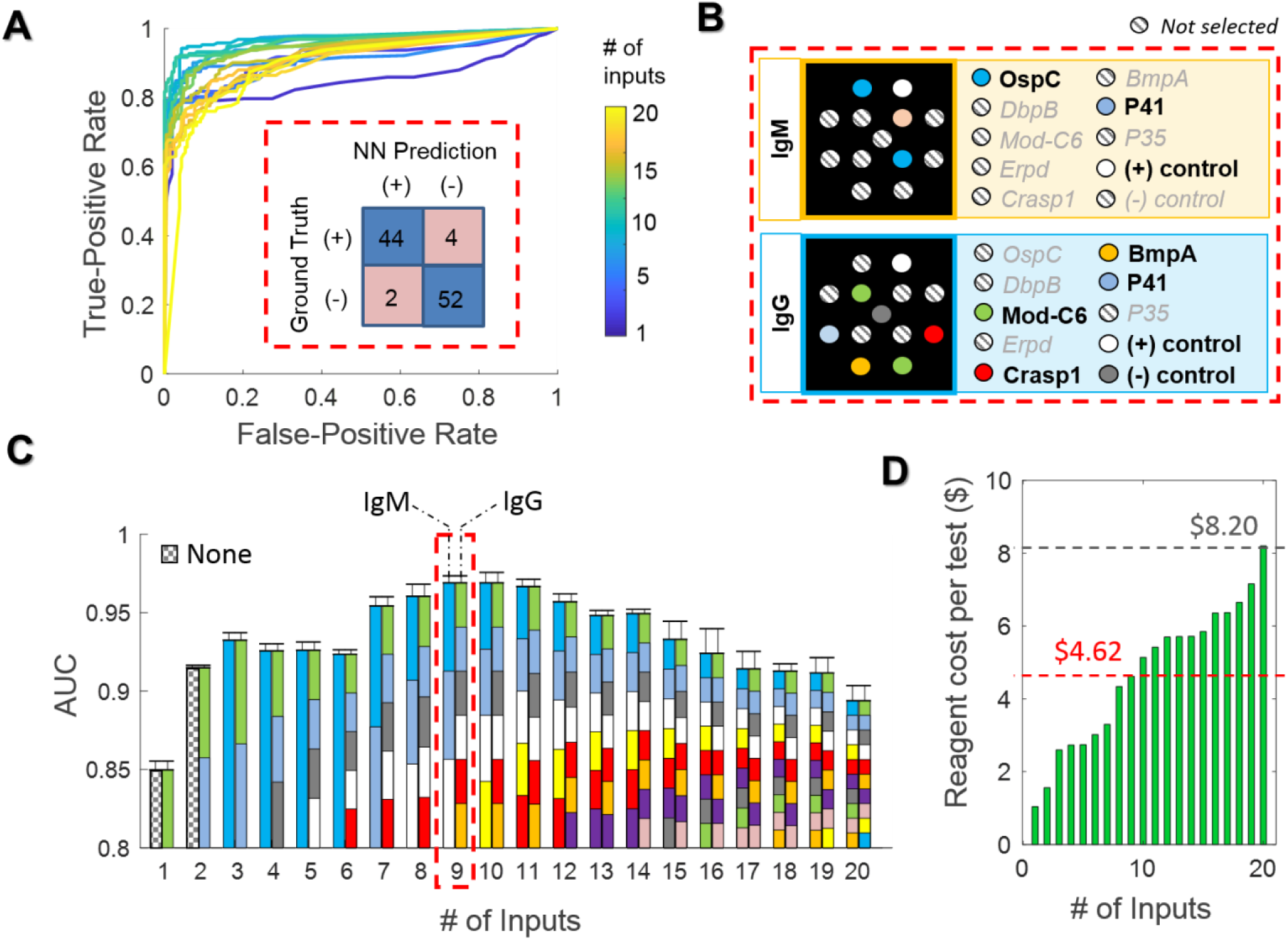
The feature selection process for the xVFA IgM and IgG antigen panel. **(A)** Receiver operator characteristic (ROC) curves resulting from the neural network inference during cross validation. The color bar represents the output from networks trained with different number of input features. The inset defined by the red dotted line shows the confusion matrix for all the samples in the training set predicted during cross-validation using the 9 antigen inputs shown in **(B)**, which is selected based on the optimization reported in **(C). (C)** The area-under-the-curve (AUC) plotted for various networks trained with a different number of inputs of the multi-antigen panel (also see **(A)**). Each bar plot corresponds to the ROC curves shown in A, and is color-coded to represent which members of the multi-antigen panel are included as the inputs with the left and right side of each bar showing the members from the IgM and IgG sensing membrane, respectively. The red-dotted line shows the local optimum AUC (0.969) and the resulting 9 selected features. The error bars show the standard deviation between 4 different training instances of the same network. **(D)** The reagent cost of the sensing membrane versus the number of selected members in the multi-antigen panel as they are included in the order shown in **(C)**. Under large-volume manufacturing the reagent cost per test is expected to drop by more than an order of magnitude. The red-dotted line represents the reagent cost for the 9 selected antigens, and the grey-dotted line shows the reagent cost of the whole antigen panel before feature selection.

Figure 4 shows the area under the receiver operator characteristic (ROC) curve (i.e., AUC) at each round of the SFFS antigen-selection algorithm, revealing a local maximum (AUC = 0.969) created by a panel of 6 antigens and the 3 control spots: OspC, P41, and the positive control spot (anti-mouse IgM) from the IgM sensing membrane, as well as the Mod-C6 peptide, Crasp1, BmpA, P41, positive control spot (anti-mouse IgG), and the negative control spot (Bovine Serum Albumin, BSA) from the IgG sensing membrane. Using this subset of sensing channels as the detection panel, a sensitivity of 91.7% and specificity of 96.2% was found via the cross-validation analysis (Fig. 4A, inset). The final LD diagnosis network was then trained using this computationally-selected subset as the antigen panel (M = 9), incorporating the tests (i.e., 100 activated xVFAs) from the first sample-pull (N_Train_ = 100, i.e., *N*_(+)_ = 48 and *N*_(−)_= 52), using the same network parameters outlined in the Methods.

#### Clinical blind testing

The second sample-pull (*N*_(+)_ = 42, *N*_(−)_= 54) was then entirely used for blind testing the performance of our xVFA diagnostic platform, yielding an AUC of 0.950, sensitivity of 90.5%, and specificity of 87.0% with respect to the seropositive and seronegative results, as summarized in Figure 5. This reported diagnostic performance of our xVFA platform achieved during the blind testing phase, to the best of our knowledge, exceeds previous POC tests for early-stage LD^31,42^. As a point of reference, Table S5 shows a comparison with the recently FDA-approved POC test from Quidel, which can be used as a first-tier test, detecting IgM and IgG antibodies using the C6 antigen. Though FDA-approved, the Quidel test is *not* recommended as a replacement of the two-tier testing. The performance of our xVFA also outperforms a number of previous clinical studies investigating diagnostic performance of standard two-tier as well as modified two-tier testing with respect to the ultimate clinical diagnosis^9,17,38^.

**Figure 5.**
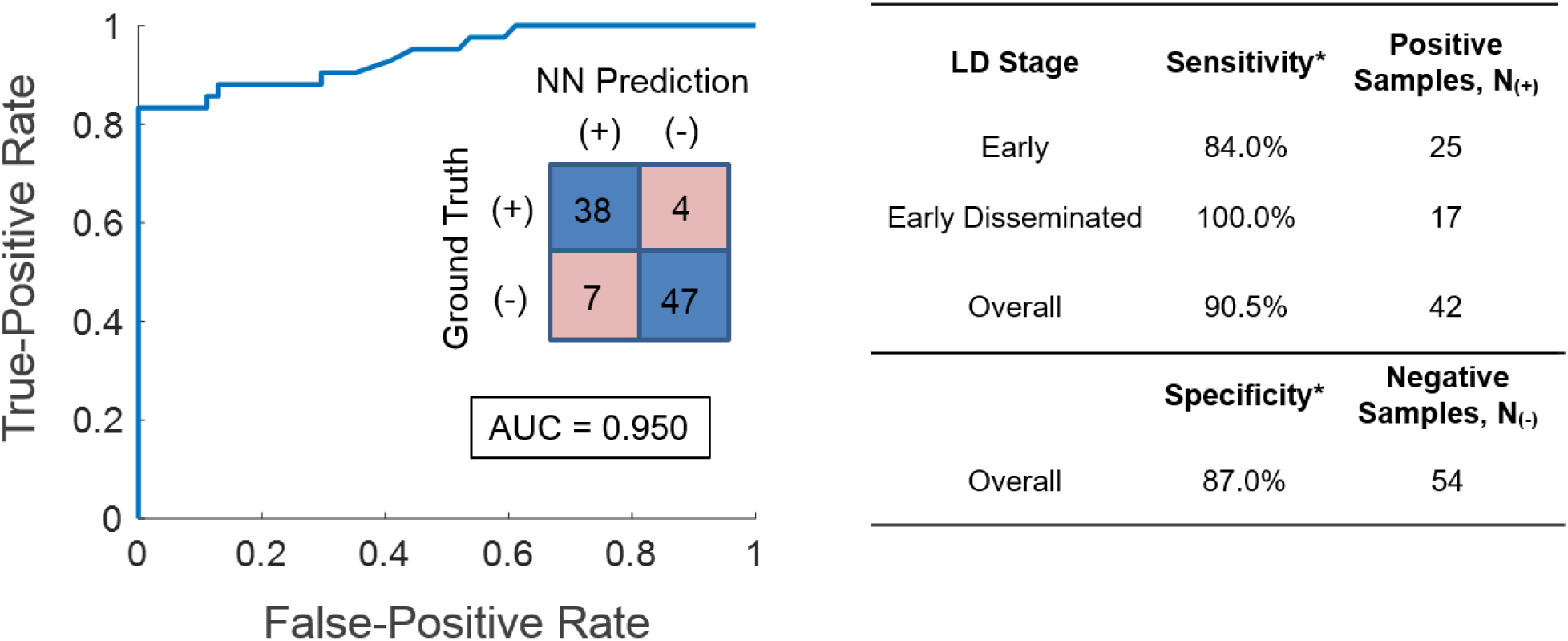
ROC curve for the blind testing data (N_Test_ = 96) as output from the neural network trained with the 9 selected antigens (see Fig. 4) from the training set (N_Train_ = 100). The inset shows the confusion matrix and area under the ROC curve (AUC). The table to the right summarizes the performance over the blindly-tested LD human serum samples with respect to the two-tier testing method.

Next, to achieve higher specificity, we tuned the decision threshold of the diagnostic network during the *training* phase such that the cross-validation specificity reached > 98%, resulting in a decision threshold of 0.66. Implementing this threshold in the blind testing phase (with 192 activated xVFAs), along with batch-specific standardization (also see the Discussion section), we achieved an AUC, sensitivity and specificity of 0.963, 85.7% and 96.3% with respect to the two-tier serology results (see Table S6). Such optimization of the decision biases can be utilized to achieve a desired false-positive and false-negative trade-off, depending on the clinical setting where the LD test is administered.

With this fine-tuned decision threshold, we achieved two false positive and six false negative results out of the 96 individual tests run through our xVFA platform, reaching an overall accuracy of 91.7%. Interestingly, some intuitive reasoning can be attributed to these instances of misdiagnosed tests. For example, four out of the six the instances of false negative tests were from two patients that self-reported the *shortest* duration of LD symptoms (≤ 1 day) indicating that these samples may have the least developed immune response in our blind testing set. Additionally, one of these two false-negative duplicate pairs (LD188, see Table S4) was clinically positive in the first-tier due only to an equivocal result in the VlsE/PepC10 EIA. Because VlsE was eliminated from our multi-antigen detection panel during the pre-screening process (see Supplementary Information, Fig. S6), our xVFA is incapable of detecting antibodies to VlsE. This can be addressed in the future by further optimizing the binding properties of VlsE antigen in the nitrocellulose substrate as well as expanding the number of diverse sera in the training set prior to the computational antigen selection.

It is also important to note that two seropositive samples in our testing set, i.e., LD185 and LD158 (see Table S4), were considered negative through standard two-tier testing (STTT), i.e. where the second-tier is IgG/IgM WB. Nevertheless, our POC test correctly called these samples positive with respect to the MTTT gold-standard label, which has demonstrated greater efficacy compared to the STTT in recent reports and has in fact lead the CDC to amend their recommendation for LD testing^16,17,38^.

Lastly, half of the misclassified samples (4 out of 8) are from single discordant tests among the duplicate pairs. Therefore, to shed more light on this result, precision testing was performed with an additional six samples (N_(+)_ = 3, N_(-)_ = 3) obtained from the LDB, where each sample was measured with six repeated xVFA tests following the same operational protocol as in the training and testing phases (see Supplementary Information, Fig. S7, S8, Table S7). To more realistically assess how the xVFA performs as a POC test, we also examined the precision between three different users/operators, two of which were new to xVFA operation and given only 5 minutes of training. With the batch-specific standardization and threshold tuning, the overall accuracy during this precision testing, 91.7%, exactly matched the accuracy obtained during the blind testing. Additionally, no difference in accuracy was observed between the precision testing from a single operator and the testing completed by the multiple newly trained operators (Table S7). The precision of the individual immunoreactions was also investigated, yielding an average coefficient of variation of 8.9% for the same operator and 9.3% for the various newly trained operators (Fig. S7, S8). While this shows good overall repeatability in the underlying immunochemistry, some antigen spots exhibited a coefficient of variation exceeding 20% which can ultimately lead to poor precision in the output of the decision algorithm. Therefore, despite the demonstration of interoperability, these testing results suggest that precision might be a limiting factor of our performance. Further improvements can be made by automating the fabrication processes and implementing humidity and temperature controlled environments, in order to improve the coefficient of variation of the individual immunoreactions as part of our xVFA. Additionally, ensuring that a large number of samples are used during the batch-specific standardization could potentially further improve our testing precision.

## DISCUSSION

### Optimization of antigen selection in early-stage LD xVFA platform

Computationally selecting the detection panel from a larger set of antigen/peptide targets improves the performance of the serodiagnostic algorithm. The capture antigens functionalized to the sensing membrane produce varying degrees of statistical variance in their optical signals, especially over different batches of fabricated sensors. This can stem from fabrication tolerances borne out of the low-cost materials, or from operational variance. Some capture antigens can also exhibit varying degrees of cross-reactivity with other antibodies native to human sera. Our feature selection procedure helps eliminate the least reliable discriminators while conserving an ensemble of reactions that can most reliably detect the immune response. A very good example of this phenomenon can be seen in this comparison: when forgoing the feature selection process and implementing the full antigen selection panel (M = 20) as inputs to the network, the cross-validation testing reveals that AUC, sensitivity and specificity are 0.894, 72.9% and 92.3% respectively, compared to 0.969, 91.7% and 96.2% respectively, from the network trained on the computationally-selected subset of 9 features (see Table S8 and Fig. 4); this clearly emphasizes that the use of more antigen-targets does not offer a better solution for LD diagnosis. Interestingly, using the single antigen with the highest t-score (the Mod-C6 like in the IgG membrane) alone as an input to the network, results in an AUC of 0.850 and a sensitivity and specificity of 77.1% and 98.1%, respectively. Such performance for early-stage LD samples is characteristic of EIAs that detect only a single antibody, as the antigen-targets employed, such as the Mod-C6, can be synthesized to limit the presence of cross-reactive epitopes while failing to detect less prevalent antibodies, such as OspC or BmpA that may be produced at the beginning of the infection, albeit at lower concentratrions^20,44^. The significant benefit of the computationally-determined multi-antigen panel is also clearly reflected in the blind testing set, showing the highest AUC when compared to networks trained with the full antigen panel and the Mod-C6 alone (see Table S8).

It is also important to note that the SFFS method implemented in this work, which is referred to as a wrapper feature selection technique, does not simply select the top individual discriminators (i.e. the sensing channels with the highest t-score). Instead, it iteratively adds input features and assesses their performance as an ensemble of inputs to a neural network. We should emphasize that filtering the input features based on the top 9 individual t-scores results in *poorer* cross-validation performance (AUC = 0.900, Sensitivity = 79.2%, Specificity = 92.3%) when compared to a network with 9 features selected by our SFFS method. This can be partly attributed to the relative unimportance of redundant information. In other words, antigen-targets associated with the same patient population or stage of infection as an already implemented antigen-target are of less value to the diagnostic performance, despite having good performance as a single discriminator; in fact, this is a *very important* conclusion for the design of multiplexed sensors in general, which certainly applies to our early-stage LD xVFA platform.

Additionally, the positive and negative control spots, while not intended for unilaterally discriminating seropositive and negative samples can contain pertinent relative information for a computational POC sensor. For example, the relative concentration of AuNPs in the conjugation pad as well as instances of strong non-specific binding from matrix effects inherent in human sera can be represented by the positive and negative control immunoreactions and thus factored into the logistical classifier.

Alternative to the wrapper SFFS technique used here, a global search could potentially be implemented in which the network is trained by *every combination* of possible sensing channels. However, this requires training a network for every possible subset of M sensing channels, multiplied by the number of folds in the k-fold cross validation, i.e. 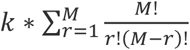. With our multiplexed xVFA platform (M = 20) and 5-fold validation, this results in over 5.2 million training instances, which was prohibitive due to the computation time required (> 10,000 hours at 7.2 seconds per training instance on an NVIDIA GeForce FTX1080 Ti GPU).

In addition to the performance advantages discussed above, our feature selection process can also be used to reduce the cost per-test. For example, the reagent cost for the full antigen panel (M=20) would be reduced by 44% by implementing the subset of 9 immunoreaction spots selected during the training phase (Fig. 4D). Although not considered here, future methodologies could even incorporate reagent cost into the feature selection cost function, *J*(***y, y***^′^) to better illustrate cost-performance trade-offs. Additionally, immunoreaction spots with antigen-targets that were not computationally selected could be replaced in future sensing membrane designs by redundancies of positive and negative control spots as well as already selected capture antigens, or even alternative antigen-targets not yet tested. This type of data-driven iterative assay development is universally applicable, and could be a powerful framework for improving multiplexed sensors, especially for complex diagnoses like LD as well as for POC tests that must balance cost and performance.

### Optimization of the false-positive and false-negative rate

Another important aspect of the training and feature selection phase is the degree of trade-off between sensitivity and specificity. The diagnostic algorithm can be influenced during the training phase by penalizing instances of false negatives more heavily than instances of false positives, e.g. through tunable weights on the two terms in the binary cross-entropy cost function used to train the network (Eq. 3), or by adjusting the threshold which discriminates between positive and negative samples. In practice, it may be more beneficial for a POC assay that is intended to be used at the first line of patient assessment, to have a greater portion of false positives over false negatives, especially in the case of contagious diseases. However, another important consideration is the pre-test likelihood, which can be low for samples submitted for serological LD testing (<20%)^24,45^. Therefore, it may also be important to ensure a lower portion of false positives in order to reduce the overall number of misdiagnoses. Ultimately however, due to the small number of misclassified samples observed during the training phase of this work (only 6), we did not implement any of these possible adjustments before the initial blind testing. With larger data-sets, where the empirical effect of tuning the bias can be better modeled, these approaches should be considered and jointly investigated with experts in diagnostic testing.

### Batch-specific standardization

Although our blind testing sensitivity is comparable to the cross-validation sensitivity during the training phase, indicating that our deep learning-based diagnostic algorithm did not over-fit to the training data-set, the network did exhibit a drift in its numerical output from the training phase to the testing set (see Fig. S9). The mean output for negative samples shifted from 0.060 ± 0.156 to 0.278 ± 0.246 (mean ± SD) between the training and testing samples (also shown in Fig. S9). This effect is likely due to the instability of the antigen-targets, which produced underlying statistical differences of the *R*_*m*_ signals between the training and testing phases of the clinical study, which were conducted 3 months apart due to sample availability from the LDB. Additionally, differences in temperature and humidity at the time of manual fabrication among other variables can impact repeatability across fabrication batches. Such issues are also pervasive in commercial assays, demanding production-grade equipment to establish environmental controls during automated fabrication as well as rigorous quality assurance to verify performance standards.

To mitigate such issues, future batches of sensors can be standardized to sample means 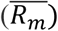 and standard deviations (*σ*_*m*_) as a characteristic signature of their fabrication batch (see Eq. 2). For example, during the initial blind testing phase reported in this work, the mean and standard deviation of the *training* data were used for the input standardization. This resulted in a drift of the *R*_*m*_′ inputs away from zero, which can in turn manifest as a drift in the network output. Therefore, by standardizing the blind testing inputs to the sample mean and standard deviation of the *testing* batch, the drift in negative sample outputs is reduced by over 80% (see Fig. S9). As a result, three of the false positive samples are subsequently classified as true negatives (with a decision threshold of 0.5), increasing the specificity from 87.0% to 92.6% and the AUC from 0.950 to 0.963 with respect to the two-tier serology, while only incurring one extra false negative sample. Practically, batch-specific sample means and standard deviations could be calculated as a running average of the activated sensors in a batch, and could even be exclusively determined using endemic control sera, which is more readily available. An alternative approach is to include fabrication batch information as inputs to the network in the form of external labels^46^. In this case, the network could learn inherent batch-to-batch variations and compensate for these differences through its internal tunable parameters.

### Outlook

In conclusion, we demonstrated a paper-based computational multi-antigen xVFA platform capable of diagnosing early-stage LD at the POC. Our xVFA has a material cost of $0.42 per-test and can be performed in 15 min by an individual with minimal training. A low-cost and handheld optical reader enables automated analysis to quantify colorimetric signals generated on a nitrocellulose membrane, followed by analysis with a neural network for inferring a diagnosis from the multi-antigen sensing information. By computationally selecting a panel of detection antigens for IgM and IgG antibodies specific to LD and performing a fully-blinded clinical study with early-stage LD samples, we report an AUC, sensitivity, and specificity of 0.950, 90.5%, and 87.0%, respectively, with respect to the two-tier serological testing. Using batch-specific standardization and threshold tuning, we improved the specificity of our blind-testing performance to 96.3%, with an AUC and sensitivity of 0.963 and 85.7%, respectively.

The multi-target and POC nature of the computational xVFA make it uniquely suited for LD diagnostics, presenting major advantages in terms of time, cost, and performance when compared to (first-tier) EIAs with single antigen-targets as well as standard two-tier testing methods that are rather costly (e.g., >$400/test) and slow (>24 hours for results).

Future work can incorporate dual-mode read-out and operation for measuring IgM and IgG antibodies in a single test. Additionally, the computational framework outlined here can be used for iteratively designing more competitive versions of our xVFA that incorporate statistically more stable, sensitive, and specific capture molecules such as synthetic peptides with epitopes engineered for high capture-affinity and low cross-reactivity^32^.

## Data Availability

We declare that all the data supporting the findings of this work are available within the manuscript. Additional data may be requested from the corresponding author.

## Acknowledgements

We acknowledge the Steven and Alexandra Cohen Foundation for their support in this study, as well as the Lyme Disease Biobank as a part of the Bay Area Lyme Foundation. The authors would also like to acknowledge Calvin Brown of UCLA Electrical and Computer Engineering for valuable discussions regarding the computational analysis.

## Notes

### Competing Interest Statement

The authors have declared no competing interest.

### Clinical Trial

Existing human serum samples were blindly obtained (without a link to the patient identifier information) from the Lyme Disease Biobank, collected under Advarra IRB# Pro00012408. These human samples were not specifically obtained for our project and were selected from existing specimen without any patient identifier information or a link to it.

### Funding Statement

We acknowledge the Steven and Alexandra Cohen Foundation for their funding support in this study.

### Author Declarations

All relevant ethical guidelines have been followed and any necessary IRB and/or ethics committee approvals have been obtained.

Any clinical trials involved have been registered with an ICMJE-approved registry such as ClinicalTrials.gov and the trial ID is included in the manuscript.

## References

1. Nelson, C. A. et al. Incidence of Clinician-Diagnosed Lyme Disease, United States, 2005–2010 - Volume 21, Number 9—September 2015 - Emerging Infectious Diseases journal - CDC. doi:10.3201/eid2109.150417

2. Stanek, G., Wormser, G. P., Gray, J. & Strle, F. Lyme borreliosis. The Lancet 379, 461–473 (2012).

3. Wormser, G. P. et al. The Clinical Assessment, Treatment, and Prevention of Lyme Disease, Human Granulocytic Anaplasmosis, and Babesiosis: Clinical Practice Guidelines by the Infectious Diseases Society of America. Clin Infect Dis 43, 1089–1134 (2006).

4. Lyme borreliosis - The Lancet. Available at: https://www.thelancet.com/journals/lancet/article/PIIS0140-6736(11)60103-7/fulltext. (Accessed: 29th August 2019)

5. Steere, A. C. & Sikand, V. K. The Presenting Manifestations of Lyme Disease and the Outcomes of Treatment. New England Journal of Medicine 348, 2472–2474 (2003).

6. Wormser, G. P. Early Lyme Disease. New England Journal of Medicine 354, 2794–2801 (2006).

7. Shapiro, E. D. Lyme Disease. New England Journal of Medicine 370, 1724–1731 (2014).

8. Adukauskienė, D. et al. Clinical relevance of high sensitivity C-reactive protein in cardiology. Medicina 52, 1–10 (2016).

9. Moore, A., Nelson, C., Molins, C., Mead, P. & Schriefer, M. Current Guidelines, Common Clinical Pitfalls, and Future Directions for Laboratory Diagnosis of Lyme Disease, United States. Emerging Infect. Dis. 22, (2016).

10. Schutzer, S. E. et al. Direct Diagnostic Tests for Lyme Disease. Clin. Infect. Dis. 68, 1052–1057 (2019).

11. Alasel, M. & Keusgen, M. Promising alternatives for one-tier testing of Lyme borreliosis. Clin. Chim. Acta 479, 148–154 (2018).

12. Dressler, F., Whalen, J. A., Reinhardt, B. N. & Steere, A. C. Western blotting in the serodiagnosis of Lyme disease. J. Infect. Dis. 167, 392–400 (1993).

13. Wormser, G. P. et al. Comparative cost-effectiveness of two-tiered testing strategies for serodiagnosis of lyme disease with noncutaneous manifestations. J. Clin. Microbiol. 51, 4045–4049 (2013).

14. Theel, E. S. The Past, Present, and (Possible) Future of Serologic Testing for Lyme Disease. J. Clin. Microbiol. 54, 1191–1196 (2016).

15. Wormser, G. P. et al. A limitation of 2-stage serological testing for Lyme disease: enzyme immunoassay and immunoblot assay are not independent tests. Clin. Infect. Dis. 30, 545–548 (2000).

16. Mead, P. Updated CDC Recommendation for Serologic Diagnosis of Lyme Disease. MMWR Morb Mortal Wkly Rep 68, (2019).

17. Marques, A. R. Revisiting the Lyme Disease Serodiagnostic Algorithm: the Momentum Gathers. J. Clin. Microbiol. 56, (2018).

18. Marques, A. R. Laboratory Diagnosis of Lyme Disease - Advances and Challenges. Infect Dis Clin North Am 29, 295–307 (2015).

19. Leeflang, M. M. G. et al. The diagnostic accuracy of serological tests for Lyme borreliosis in Europe: a systematic review and meta-analysis. BMC Infectious Diseases 16, 140 (2016).

20. Nowalk, A. J., Gilmore, R. D. & Carroll, J. A. Serologic Proteome Analysis of Borrelia burgdorferi Membrane-Associated Proteins. Infection and Immunity 74, 3864–3873 (2006).

21. Magnarelli, L. A., Ijdo, J. W., Padula, S. J., Flavell, R. A. & Fikrig, E. Serologic diagnosis of Lyme borreliosis by using enzyme-linked immunosorbent assays with recombinant antigens. J. Clin. Microbiol. 38, 1735–1739 (2000).

22. Lahdenne, P. et al. Improved serodiagnosis of erythema migrans using novel recombinant borrelial BBK32 antigens. J. Med. Microbiol. 52, 563–567 (2003).

23. Engstrom, S. M., Shoop, E. & Johnson, R. C. Immunoblot interpretation criteria for serodiagnosis of early Lyme disease. J. Clin. Microbiol. 33, 419–427 (1995).

24. Hinckley, A. F. et al. Lyme Disease Testing by Large Commercial Laboratories in the United States. Clin Infect Dis 59, 676–681 (2014).

25. Gomes-Solecki, M. J. C. et al. A First-Tier Rapid Assay for the Serodiagnosis of Borrelia burgdorferi Infection. Arch Intern Med 161, 2015–2020 (2001).

26. López-Marzo, A. M. & Merkoçi, A. Paper-based sensors and assays: a success of the engineering design and the convergence of knowledge areas. Lab Chip 16, 3150–3176 (2016).

27. Smith, S., Korvink, J. G., Mager, D. & Land, K. The potential of paper-based diagnostics to meet the ASSURED criteria. RSC Adv. 8, 34012–34034 (2018).

28. Quidel Receives FDA Clearance, CLIA Waiver for Its Point-of-Care Sofia® 2 Lyme Fluorescent Immunoassay for Use with Sofia® 2 Instrument from Finger-Stick Whole Blood Specimens. Quidel Corporation Available at: http://ir.quidel.com/news-releases/news-release-details/quidel-receives-fda-clearance-clia-waiver-its-point-care-sofiar. (Accessed: 3rd September 2019)

29. Liang, F. T. et al. Sensitive and Specific Serodiagnosis of Lyme Disease by Enzyme-Linked Immunosorbent Assay with a Peptide Based on an Immunodominant Conserved Region of Borrelia burgdorferi VlsE. Journal of Clinical Microbiology 37, 3990–3996 (1999).

30. Arnaboldi, P. M. & Dattwyler, R. J. Cross-Reactive Epitopes in Borrelia burgdorferi p66. Clin. Vaccine Immunol. 22, 840–843 (2015).

31. Maulden, A. B. et al. Two-Tier Lyme Disease Serology Test Results Can Vary According to the Specific First-Tier Test Used. J Pediatric Infect Dis Soc (2019). doi:10.1093/jpids/piy133

32. Lahey, L. J. et al. Development of a Multiantigen Panel for Improved Detection of Borrelia burgdorferi Infection in Early Lyme Disease. J. Clin. Microbiol. 53, 3834–3841 (2015).

33. Barbour, A. G. et al. A genome-wide proteome array reveals a limited set of immunogens in natural infections of humans and white-footed mice with Borrelia burgdorferi. Infect. Immun. 76, 3374–3389 (2008).

34. Porwancher, R. B. et al. Multiplex immunoassay for Lyme disease using VlsE1-IgG and pepC10-IgM antibodies: improving test performance through bioinformatics. Clin. Vaccine Immunol. 18, 851–859 (2011).

35. Bejnordi, B. E. et al. Diagnostic Assessment of Deep Learning Algorithms for Detection of Lymph Node Metastases in Women With Breast Cancer. JAMA 318, 2199–2210 (2017).

36. Esteva, A. et al. A guide to deep learning in healthcare. Nat Med 25, 24–29 (2019).

37. Hu, L. et al. An Observational Study of Deep Learning and Automated Evaluation of Cervical Images for Cancer Screening. J Natl Cancer Inst doi:10.1093/jnci/djy225

38. Branda, J. A. et al. Advances in Serodiagnostic Testing for Lyme Disease Are at Hand. Clin. Infect. Dis. 66, 1133–1139 (2018).

39. Bouchard, C. et al. The increasing risk of Lyme disease in Canada. Can Vet J 56, 693–699 (2015).

40. Hofhuis, A., Harms, M., van den Wijngaard, C., Sprong, H. & van Pelt, W. Continuing increase of tick bites and Lyme disease between 1994 and 2009. Ticks and Tick-borne Diseases 6, 69–74 (2015).

41. Geographic Distribution and Expansion of Human Lyme Disease, United States. Emerg Infect Dis 21, 1455–1457 (2015).

42. Nayak, S. et al. Microfluidics-based point-of-care test for serodiagnosis of Lyme Disease. Scientific Reports 6, 35069 (2016).

43. Joung, H.-A. et al. Paper-based multiplexed vertical flow assay for point-of-care testing. Lab Chip (2019). doi:10.1039/C9LC00011A

44. Gomes-Solecki, M. J. C. et al. Recombinant Chimeric Borrelia Proteins for Diagnosis of Lyme Disease. Journal of Clinical Microbiology 38, 2530–2535 (2000).

45. DePietropaolo, D. L., Powers, J. H., Gill, J. M. & Foy, A. Diagnosis of Lyme Disease. AFP 72, 297–304 (2005).

46. Ballard, Z. et al. Deep Learning-Enabled Point-of-Care Sensing Using Multiplexed Paper-Based Sensors. bioRxiv 667436 (2019). doi:10.1101/667436

